# The *p* Factor Outweighs the Specific Internalizing Factor in Predicting Recurrences of Adolescent Depression

**DOI:** 10.1101/2023.08.11.23293988

**Authors:** Yinuo Shu, Na Ao, Xue Wen, Zaixu Cui, Diyang Qu, Runsen Chen

## Abstract

**Background:** The early prediction of adolescent depression recurrence poses a significant challenge in the field. This study aims to investigate and compare the abilities of two psychopathology factors, namely the general psychopathology factor (*p*) and the specific internalizing factor, in predicting depression recurrence over a 2-year course, as well as identifying remitted depression patients from healthy adolescents. Longitudinal changes of these two factors in different trajectory groups were also tracked to examine their sensitivity to sustained remission and relapse.

**Methods:** We included 255 baseline-remitted depression patients and a healthy control group (n=255) matched in age, sex, and race, sourced from the Adolescent Brain Cognitive Development Study. The Linear Mixed Model was employed to differentiate between healthy controls and remitted depression patients, predict depression recurrence, and track longitudinal changes over a subsequent 2-year course using the *p* factor and the specific internalizing factor respectively.

**Results:** The *p* factor not only effectively discriminated between remitted depression patients and healthy controls, but also robustly predicted the depression recurrence over a subsequent 2-year course. The specific internalizing factor could only differentiate remitted depression patients from healthy controls. Additionally, a noteworthy longitudinal decline of the *p* factor in the sustained-remission group was observed.

**Conclusions:** Psychopathology factors serve as the inherent and enduring measurement of long-term mental health aberrations. Longitudinal results indicate that the p factor is more sensitive to respond to sustained remission than the internalizing factor. The ability of the overall *p* factor to anticipate depression relapse, unlike the specific internalizing factor, suggests clinical interventions should monitor and mitigate the coincident symptoms across all dimensions to preempt relapse of adolescent depression, rather than an exclusive focus on internalizing symptoms.

## Introduction

Depressive disorders, as an umbrella term, ranging from major depression to atypical depression to dysthymia, has become one of the most serious mental health concerns and leading contributors of the global health-related burden nowadays (Smith, 2014; Friedrich, 2017; Herrman et al., 2022; Marcus et al., 2012). Adolescents are particularly at risk for developing depression, with estimates of major depressive disorder ranging from 8% to 20% occurring before the age of 18 (Cheung & Dewa, 2006; Hankin et al., n.d.; Kessler & Walters, 1998; Naicker et al., 2013). Notably, depression originating in adolescence frequently manifests in a recurring manner and tends to be associated with more severe outcomes compared to adult-onset depression (Birmaher, 2002; Fombonne et al., 2001; Lewinsohn et al., 1999; Weissman, 1999). This recurrent pattern can lead to substantial impairments across crucial psychosocial domains, with effects that may persist into adulthood (Brière et al., 2014; Clayborne et al., 2019; Johnson et al., 2018; Wilson & Dumornay, 2022). However, there is no permanent treatment solution for depressive disorders due to their relapsing-remitting nature. Individual experience a relapse after the treatment of their first episode of depression may tend to recur with greater severity and with lessening responsivity to conventional treatments (Ali et al., 2017). Therefore, there is a pressing need for a deeper understanding of early-stage markers that can predict the later development of depression recurrence (Burcusa & Iacono, 2007; Colizzi et al., 2020; Davey & McGorry, 2019; Johnson et al., 2018). Monitoring changes in markers can help individuals and healthcare professionals become aware of early warning signs of a potential relapse. This awareness allows them to implement tailored interventions, enhancing the effectiveness of relapse prevention and even circumventing the onset of initial treatment resistance. In another word, this approach may heighten the prospect of sustained recovery and prevent the intensification of depression during the later stages of adulthood (Dinga et al., 2018).

Prior research has identified various risk factors for depression recurrence in remitted patients, including a higher number of preceding episodes, higher levels of residual symptoms, lower levels of positive refocusing (ten Doesschate et al., 2010), presence of anxiety (Penninx et al., 2011), longer symptom duration, higher symptom severity, and earlier age of onset (Pettit et al., 2009). While previous studies have made valuable contributions to our understanding of predictors for depression recurrence, a shared limitation is their focus on syndrome-specific indicators such as the Patient Health Questionnaire-9 (PHQ-9) or the Hamilton Depression Rating Scale (HAMD), designed to detect specific signs or symptoms of depression. However, considering the latent intricate etiology of depression, which encompasses interplays of a broader spectrum of symptoms across multiple dimensions and a high degree of comorbidity with other disorders like externalizing disorders that can significantly impact the trajectory of depression and increase the risk of further recurrence (Wolff & Ollendick, 2006), the incorporation of multi-dimensional psychopathology becomes a necessity. Therefore, using the indicators that encompasses information from symptoms across multiple dimensions could potentially yield a more accurate prediction of depression recurrence. Recent advancements in psychopathology studies have indicated an overall latent factor, the general psychopathology factor, that may further provide an explanation for these pathways (Smith et al., 2020).

The general psychopathology factor, commonly referred to as *p* factor, accounts for common variance across a wide range of symptoms spanning multiple diagnostic domains (Murray et al., 2016). It embodies shared aspects among various mental disorders (Patalay et al., 2015), and directly impacts symptoms across distinct dimensions (Caspi et al., 2014). Previous research has identified the presence of the *p* factor in adolescents and suggested that investigating this factor could enhance our understanding of the etiology, risk, and correlates of psychopathology in this age group (Patalay et al., 2015; Sallis et al., 2019; Laceulle et al., 2015). For instance, Moore et al. (Moore et al., 2020) identified the overall *p* factor through a bifactor model using Child Behavior Checklist (CBCL) from the Adolescent Brain Cognitive Development (ABCD) Study. In this bifactor model, three lower-level factors in distinct domains (Internalizing, Attention Deficit Hyperactivity Disorder (ADHD), and Conduct Problems) have also been identified, that accounts for shared variance within a specific dimension, from which the overall variance (*p*) across all dimensions has been subtracted.

Indeed, studies have suggested that the *p* factor in adolescents may be more predictive of long-term adverse mental health outcomes, including diagnoses of depression and anxiety, psychological well-being, criminal activity, alcohol use, and educational attainment, compared to syndrome-specific factors. These findings indicate that interventions should focus on addressing the co-occurrence of internalizing and externalizing symptoms to mitigate the long-term impact on individuals (Sallis et al., 2019).

However, there is limited knowledge regarding the prognostic effect of the overall *p* factor or the specific lower-level internalizing factor to which depression is directly related, on the prediction of adolescent depression recurrences. Additionally, the comparative performance of these two factors in terms of their predictive ability in depression recurrence remains unknown. Further research is needed to understand the specific contributions of the internalizing factor and the *p* factor in predicting depression trajectories and to compare their predictive abilities. This information would be valuable in guiding interventions and improving the effectiveness of targeted treatments for depression in adolescents.

To fill this research gap, our study evaluated the capacity of the two factors—the *p* factor and the specific internalizing factor—all measured during a remitted state, to predict the recurrence of depression over a 2-year period, and to discriminate between depression patients and their healthy counterparts. This was achieved by using two waves of clinical data collected at baseline and at a 2-year-follow-up from the ABCD Study. Simultaneously, we tracked the longitudinal change of each factor over 2 years to detect their sensitivity in response to either sustained remission or relapse. Our aim is to enhance our understanding of the underlying mechanisms involved in the recurrence of adolescent depression and provide valuable insights into effective intervention strategies for managing recurrent depression in adolescents.

## Methods

### Participants

Data for this study were derived from a large-scale, multi-site, and longitudinal study in the United States: the Adolescent Brain Cognitive Development (ABCD) Study® (Release 3.0, November 2020) (Casey et al., 2018). This extensive dataset included comprehensive clinical, behavioral, cognitive, and multi-modal neuro-imaging data collected at four distinct timepoints (baseline, 1-year follow-up, 2-year follow-up, and 3-year follow-up). The current research focused on a portion of the baseline data (n=11,876, aged 9-10 years) and the 2-year-follow-up data (n=10,404, aged 11-12 years) within the ABCD Study, given that Kiddie Schedule for Affective Disorders and Schizophrenia (KSADS) depressive diagnostic information was collected biennially.

Depressive disorder diagnoses were determined using parent or guardian ratings in the computerized KSADS based on Diagnostic and Statistical Manual of Mental Disorders, Fifth Edition (DSM-V) criteria (Barch et al., 2018). Our study included 255 subjects who met our selection criteria (Fig. 1): (i) presence of a diagnosed past Major Depressive Disorder (MDD), Dysthymia, or unspecified depressive disorder at baseline; (ii) exclusion of a diagnosed bipolar disorder, psychosis, or substance use at either baseline or 2-year-follow-up. It should be noted that our study concentrated on subjects who received KSADS diagnoses of past (in a remitted state at the moment of baseline measurement) depressive disorders at the baseline. This was due to the limited number of subjects diagnosed with present depressive disorders at both timepoints.

**Figure 1.**
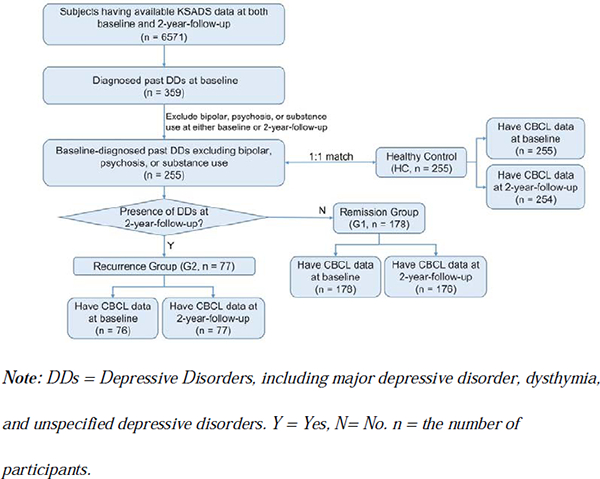
Flowchart of participant selection criteria and group allocation.

The current study also involved a control group of 255 healthy individuals (HC, M_age_ = 118.41, SD = 7.20; 53.73% were girls, and 50.98% were non-Hispanic White) who showed none of any KSADS diagnoses at both the baseline and the 2-year follow-up. These control subjects were matched with the 255 depression patients on age, sex, and race. We have also tested all subsequent statistical analyses utilizing the unmatched healthy control sample (n = 1597), and the outcomes aligned perfectly with those derived from the matched healthy controls. We chose to present the results associated with the matched healthy controls in the main body of our text, while the outcomes derived from the unmatched healthy control sample are included in the appendix for comprehensive review and comparison (Supplementary Table 2, Supplementary Figure 1).

### Definition of diagnosis trajectory groups

We defined the diagnosis trajectory groups according to the absence (G1, n=178; M_age_ =119.23, SD = 7.24; 44.38% were girls and 51.69% were non-Hispanic White) or presence (G2, n=77; M_age_ =121.38, SD = 7.74; 49.35% were girls and 61.04% were non-Hispanic White) of 2-year-follow-up parent-report KSADS diagnosed MDD, Dysthymia, or unspecified depressive disorder that was present (in the recent two weeks, n=4), in partial remission (n=4), or past (since baseline assessment, n=75) (Fig. 1). According to the defined criteria, G1 represented a remission group with participants who experienced no recurrence of depression for a minimum of 2 years; while G2 represented a recurrence group, consisting of individuals who were in remission at the baseline measurement but experienced a recurrence over the subsequent two-year period. Thus, G1 demonstrated a more favorable trajectory, tending towards stable remission, while G2 exhibited a recurrence of depression within the 2-year course.

### Demographic information

Age, sex, race, site was included as covariates. The specific files from which these demographic variables were derived were presented in the Supplementary Table 1.

### Measure of psychopathology

Psychopathology was measured from parent-reported Child Behavior Checklist (CBCL) which was used to assess emotional and behavioral problems in school-aged children (Achenbach, 2015). All 119 items were scored using a 3-point Likert scale, ranging from 0 (“not true”) to 2 (“very true”). As previously described (Moore et al., 2020), four-dimensional psychopathology factors (including the general psychopathology factor, or *p* factor, and three lower-level factors: internalizing, ADHD, and conduct problems) were derived employing a bifactor model, which has demonstrated robust reliability and validity. The final model included 66 items from which the *p* factor and internalizing factor scores were computed for each participant at both baseline and 2-year-follow-up. For further information regarding the calculation procedures and results of the bifactor modeling, as well as the validity and reliability of the psychopathology dimensions, please refer to Moore et al. (2020).

### Statistical Analysis

#### Predict depression trajectories

Generalized Linear Mixed Model (GLMM) were applied to predict depression trajectory groups utilizing R lmeTest packages (Kuznetsova et al., 2017). For the two trajectory groups (G1, G2), the ability of the baseline *p* factor and baseline specific internalizing factor respectively in predicting trajectory groups were examined. In the GLMM formula, the *p* factor and the specific internalizing factor served as the independent variable respectively, the group was the dependent variable, and age, sex, and race were employed as fixed-effects covariates, while site was utilized as a random-effects covariate (Brooks et al., 2017). All the continuous variables were standardized, including the *p* factor, the specific internalizing factor, and age. Other categorical variables were dummy coded before being put into the model, including group, sex, race, and site (Wen et al., 2023). False Discovery Rate (FDR) was applied for multiple comparisons to avoid the type I errors.

#### Distinguish remitted depression patients from healthy controls

The same GLMM model was used to distinguish between depression patients and healthy controls, only differing in that the dependent variable “group” was either HC and G1 or HC and G2. FDR was applied for multiple comparisons.

#### Longitudinal analysis

The longitudinal alterations of the *p* factor and the specific internalizing factor in each group were investigated by employing Linear Mixed Model (LMM). For each group, the *p* factor and the specific internalizing factor functioned as the dependent variable respectively, the time variable (baseline defined as 0, 2-year follow-up defined as 1) served as the independent variable, and sex and race were used as fixed-effects covariates, with site as a random-effects covariate. Additionally, the subject was incorporated as a random-effects covariate to eliminate individual differences. FDR was applied for multiple comparisons.

## Results

### Psychopathology Factors Distinguish Remitted Depression Patients from Healthy Controls

Both the *p* factor and the internalizing factor were capable of distinguishing depression patients from healthy controls, as both G1 and G2 exhibited significantly higher *p* factor than HC at both baseline (G1 vs. HC: p_FDR_ = 2.95×10^-23^; G2 vs. HC: p_FDR_ = 6.11×10^-15^) and 2-year-follow-up (G1 vs. HC: p_FDR_ = 2.23×10^-20^; G2 vs. HC: p_FDR_ = 1.66×10^-14^) (Fig. 2 A) as well as higher internalizing factor than HC at both baseline (G1 vs. HC: p_FDR_ = 2.22 × 10^-9^; G2 vs. HC: p_FDR_ = 1.06× 10^-8^) and 2-year-follow-up (G1 vs. HC: p_FDR_ = 3.11×10^-7^; G2 vs. HC: p_FDR_ = 1.06×10^-11^). The boxplots of the *p* factor and the internalizing factor in each subgroup at baseline and 2-year-follow-up were presented in Fig. 2 B. Detailed modelling results were showed in the Table 1.

**Figure 2.**
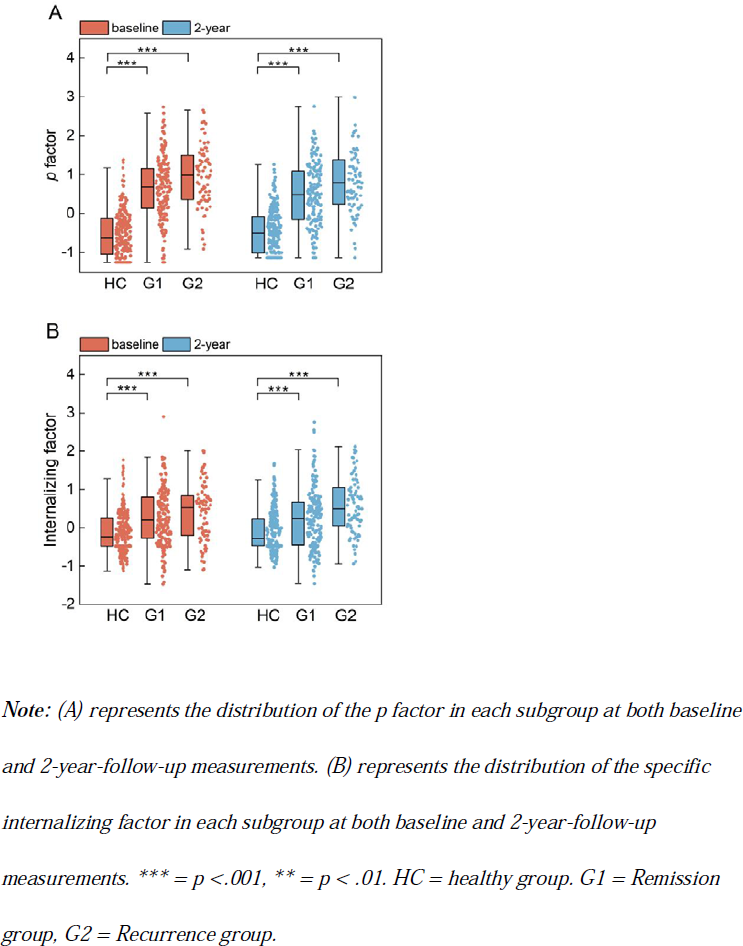
Results of the *p* factor and internalizing factor distinguishing between remitted depression patients and healthy controls at baseline and 2-year-follow-up.

**Table 1.**
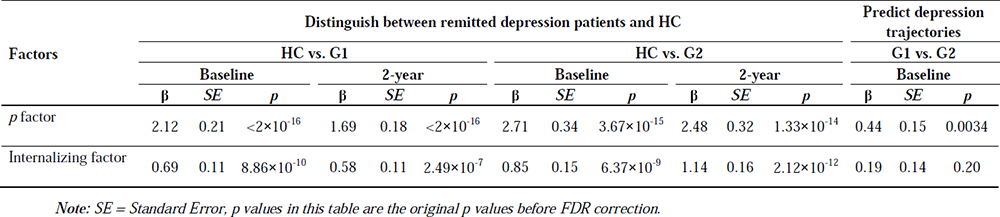
Generalized Linear Mixed Modelling results after controlling for age, sex, race, and site.

### The *p* Factor Predicts Depression Trajectories

Of the two factors examined, only the baseline *p* factor was found to be capable of predicting depression trajectories over the subsequent 2-year period (p_FDR_ = 0.0034), with a higher *p* factor being indicative of depression recurrence (Fig. 3 A). However, the baseline internalizing factor failed to exhibit predictive ability in predicting depression trajectories over the next 2 years. Detailed modelling results were showed in the Table 1.

**Figure 3.**
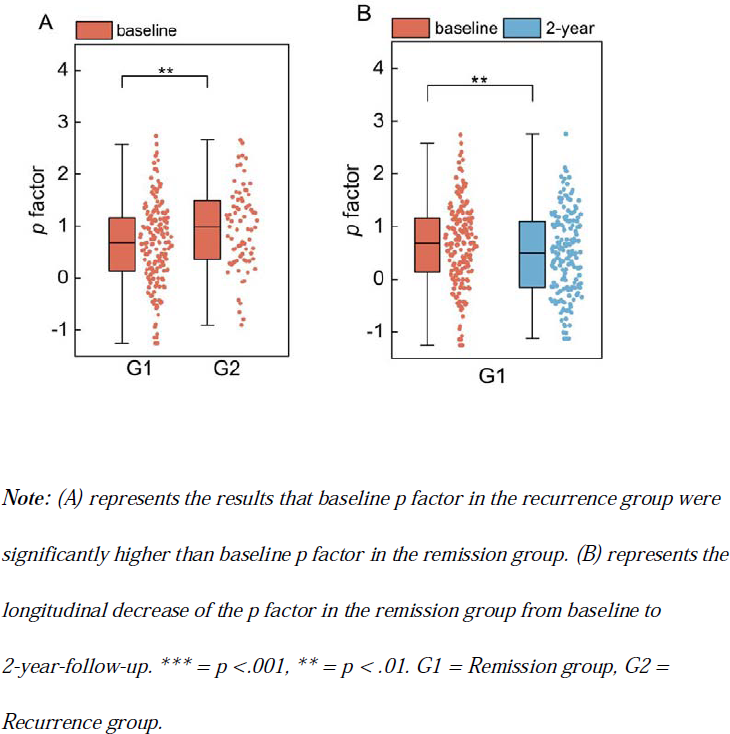
Results of the *p* factor predicting depression trajectories and mirroring sustained remission over a 2-year course.

### Longitudinal Decrease of the *p* Factor in the Remission Group

Among the three groups, only G1 exhibited a significant decrease in *p* factor from baseline to 2-year-follow-up (p_FDR_ = 0.0041) (Fig. 3 B). And no significant changes of the specific internalizing factor were found in any of the three groups. Detailed modelling results were showed in the Table 2.

**Table 2.**
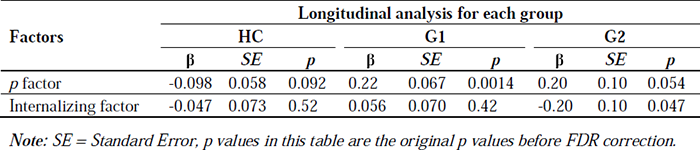
Linear Mixed Modelling results of longitudinal analysis after controlling for age, sex, race, and site.

## Discussion

The effective tool for predicting the developmental trajectories, especially the relapse of adolescent depression, is critical for tailing treatments and interventions for adolescent depression recurrence. Little is known about the extent to which the latent psychopathology factors contribute to predicting the recurrence of adolescent depression. This study aims to examine and compare the impact of the specific internalizing factor and the *p* factor on the prediction of depression recurrence, as well as their ability to differentiate between currently remitted depression patients and healthy individuals. Our research findings provide valuable clinical insights. For instance, both the specific internalizing factor and the *p* factor contribute to distinguishing adolescents with a history of depression from healthy populations. However, it is noteworthy that only the *p* factor demonstrates its sensitivity as an effective tool in predicting later depression relapse. Additionally, there is a consistent decline of the *p* factor in sustained remission patients over time, which can be used as an indicator of long-term stable remission. Taken together, our findings reveal that latent psychopathology factors may serve as inherent and enduring indicators for long-term mental health aberrations, while the *p* factor exhibits sensitivity to both relapse and sustained remission of depression. Therefore, it is crucial to emphasize the importance of monitoring and intervening in the co-occurrence of symptoms across all dimensions, represented by the *p* factor, as an effective tool to prevent the recurrence of adolescent depression.

We firstly assessed the ability of the specific internalizing factor and *p* factor respectively in distinguishing currently remitted depression patients from their healthy counterparts. Our results revealed that both the specific internalizing factor and *p* factor were found to be significantly higher in depression patients compared to healthy controls even if measured at a remitted state. This observation indicates that both the overall the specific internalizing factor and *p* factor are capable of capturing residual symptoms in remitted depression patients, therefore, greatly equipped to identifying remitted depression patients. And this finding is also consistent with views common in the antidepressant field that the depressive disorder is merely suppressed at a remitted state and the underlying disturbance continues until spontaneous remission occurs (Paykel, 2008), which explains why the remitted patients exhibited higher specific internalizing factor and *p* factor than their healthy counterparts.

Subsequently, we examined the prognostic effect of the two factors on depression recurrence separately. Our study specifically found that an elevated baseline *p* factor (not the internalizing factor), measured during a remitted state, can effectively predict the recurrence of depression in the following two years. This observation aligns with a recent study that discovered patients with a high baseline *p* factor were more likely to experience poorer outcomes in terms of short-term psychotherapy response (Fiorini et al., 2023). This observation reflects the relatively insensitive or insubstantial role of the specific internalizing factor in predicting depression recurrence. In contradistinction, the overarching *p* factor presents itself as a more sensitive predictor, aligning with prior research that indicates the *p* factor’s greater relevance for long-term outcomes compared to specific factors (Sallis et al., 2019).

It is important to note that our findings do not imply that internalizing symptoms have no association with future depression recurrence. In fact, our results suggest that once the shared variance across all dimensions (the *p* factor) is taken into account, the remaining unique variance (the specific internalizing factor) does not significantly relate to future depression recurrence (Sallis et al., 2019). This observation highlights the crucial role of symptoms in other dimensions of psychopathology, such as ADHD and Conduct Problems, in the development of future episodes. In line with the transdiagnostic approach, the findings lead us to consider recurrent depression as potentially arising from complex interactions among symptoms across all dimensions, rather than being strictly confined to the internalizing dimension(Lilienfeld, 2003; Wolff & Ollendick, 2006). By acknowledging the intricate interplay of symptoms across diverse dimensions, we gain a deeper understanding of the complex nature of recurrent depression and its underlying mechanisms. This realization prompts a shift in perspective, which highlighting the importance of monitoring and addressing comorbid symptoms across all dimensions (Dalgleish et al., 2020). In this process, the *p* factor emerges as a highly sensitive tool for detecting the risk of future depression recurrences (Caspi et al., 2014). Further research is warranted to delve deeper into the mechanisms underlying the influence of the specific psychopathology factors, especially given that the total variance of the *p* factor has been accounted for (Smith et al., 2020).

Furthermore, even after an extended period of remission (at least two years), both the specific internalizing factor and the *p* factor in the remission group remained considerably higher than that of the healthy controls at the 2-year-follow-up measurement. This observation suggests that individuals with a history of depression might continue to exhibit a higher specific internalizing factor and *p* factor compared to those without such a history, even during prolonged, stable recovery and when deemed healthy at the time of assessment. This observation indicates that latent psychopathology factors reflect inherent and enduring mental health deviations, which may serve as a straightforward and effective measurement for lifetime psychopathology evaluations.

Interesting, the longitudinal analysis over a 2-year period showed a significant decrease in the *p* factor within the remission group. However, no significant longitudinal changes were found in the specific internalizing factor. This observation aligns with a previous study which observed a diminishing pattern in the *p* factor during short-term psychotherapies, while the specific lower-level factors remained stable (Fiorini et al., 2023). This finding suggests that the sustained depression remission may not necessarily induce significant changes in the specific characteristics of internalizing dimension when *p*’s variance was accounted for. In contrast, the fluctuation of *p* factor is more sensitive to reflect sustained remission. Therefore, the significant decline in the *p* factor could be considered as a more sensitive and effective indicator for detecting the long-term remission of depression compared to the lower-level specific internalizing factor.

Several limitations inherent to our study should be acknowledged. Firstly, the delineation of depression trajectories in our study was based on KSADS diagnoses at two timepoints. While informative, this approach may not capture the full nuance of depression trajectories. Future research should aim to explore more precisely defined depression trajectories. Secondly, we were unable to consider factors such as first onset age, number of episodes, and antidepressant treatment due to the lack of available data from the ABCD cohort. It would be valuable for subsequent studies to explore the influence of these factors on predicting depression recurrence. Despite these limitations, our study provides valuable insights and lays the groundwork for further research in this area.

## Conclusion

In conclusion, our study shed light on the critical role of the *p* factor, rather than the specific internalizing factor, in predicting future recurrence of adolescent depression and mirroring sustained remission. Moreover, our study suggested the importance of monitoring and intervening co-occurrence of symptoms across all dimensions in preventing adolescent depression recurrence, rather than solely focusing on the internalizing dimension. Further research examining the *p* factor in predicting adolescent depression trajectories over an extended period and investigating novel interventions and treatments aimed at mitigating symptoms across all dimensions and reducing the *p* factor could be conducted.

## Data Availability

All data produced in the present study are available upon reasonable request to the authors.
All data produced are available online at https://nda.nih.gov/general-query.html?q=query=featured-datasets:Adolescent%20Brain%20Cognitive%20Development%20Study%20(ABCD)

## Authors’ Contributions

Yinuo Shu was responsible for the study conception and design, statistical analysis, first draft writing and revision. Na Ao was responsible for validating the results. Xue Wen was responsible for calculating the psychopathology factors. Zaixu Cui, Diyang Qu, and Runsen Chen were responsible for mentoring the whole study and revising the manuscript. All authors commented on previous versions of the manuscript. All authors read and approved the final manuscript.

## Acknowledgements

We thank the Adolescent Brain Cognitive Development (ABCD) participants and their families for their time and dedication to this project. Data used in the preparation of this article were obtained from the ABCD Study (https://abcdstudy.org) and are held in the NIMH Data Archive (NDA). This is a multisite, longitudinal study designed to recruit more than 10,000 children aged 9–10 and follow them over 10 years into early adulthood. The ABCD Study is supported by the National Institutes of Health (NIH) and additional federal partners under award numbers U01DA0401048, U01DA050989, U01DA051016, U01DA041022, U01DA051018, U01DA051037, U01DA050987, U01DA041174, U01DA041106, U01DA041117, U01DA041028, U01DA041134, U01DA050988, U01DA051039, U01DA041156, U01DA041025, U01DA041120, U01DA051038, U01DA041148, U01DA041093, U01DA041089, U24DA041123, and U24DA041147. A full list of supporters is available at https://abcdstudy.org/federal-partners.html. A listing of participating sites and a complete listing of the study investigators can be found at https://abcdstudy.org/principal-investigators/. ABCD consortium investigators designed and implemented the study and/or provided data but did not necessarily participate in the analyses or writing of this report. This manuscript reflects the views of the authors and may not reflect the opinions or views of the NIH or ABCD consortium investigators.

