## Supplementary Table 1 for "The *p* Factor Outweighs the Specific Internalizing Factor in Predicting Recurrences of Adolescent Depression"

### SUPPLEMENTARY MATERIALS

**Supplementary Table 1.** *ABCD data release 3.0 variables used in current analysis*

| Variable labels in current report | Variable labels in dataset | Scales in dataset |
|---|---|---|
| Sex | sex | pdem02 |
| Age | interview_age | pdem02 |
| Site | site_id_l | abcd_lt01 |
| Race/Ethnicity | race_ethnicity | acspsw03 |

**Supplementary Table 2.** *Generalized Linear Mixed Modelling results using full healthy control sample (n = 1597) after controlling for age, sex, race, and site*

| Metrics | Distinguish between remitted depression patients and HC | | | | | | | | | | | | | | | |
|---|---|---|---|---|---|---|---|---|---|---|---|---|---|---|---|---|
| | HC vs. G1 | | | | | | | | HC vs. G2 | | | | | | | |
| | Baseline | | | | 2-year | | | | Baseline | | | | 2-year | | | |
| | β | *SE* | *p* | $p_{FDR}$ | β | *SE* | *p* | $p_{FDR}$ | β | *SE* | *p* | $p_{FDR}$ | β | *SE* | *p* | $p_{FDR}$ |
| P factor | 1.71 | 0.11 | $<2\times10^{-16}$ | $7.66\times10^{-52}$ | 1.37 | 0.096 | $<2\times10^{-16}$ | $3.63\times10^{-46}$ | 2.00 | 0.18 | $<2\times10^{-16}$ | $1.48\times10^{-29}$ | 1.75 | 0.15 | $<2\times10^{-16}$ | $7.45\times10^{-31}$ |
| Internalizing factor | 0.57 | 0.074 | $8.35\times10^{-15}$ | $2.51\times10^{-14}$ | 0.55 | 0.074 | $1.20\times10^{-13}$ | $2.41\times10^{-13}$ | 0.71 | 0.10 | $8.97\times10^{-12}$ | $1.35\times10^{-11}$ | 0.98 | 0.11 | $<2\times10^{-16}$ | $9.83\times10^{-19}$ |

***Note:*** *SE = Standard Error, p values in this table are the original p values before FDR correction.*

**Supplementary Figure 1.** *Results of the p factor and the internalizing factor distinguishing between remitted depression patients and full sample healthy controls (n = 1597)*

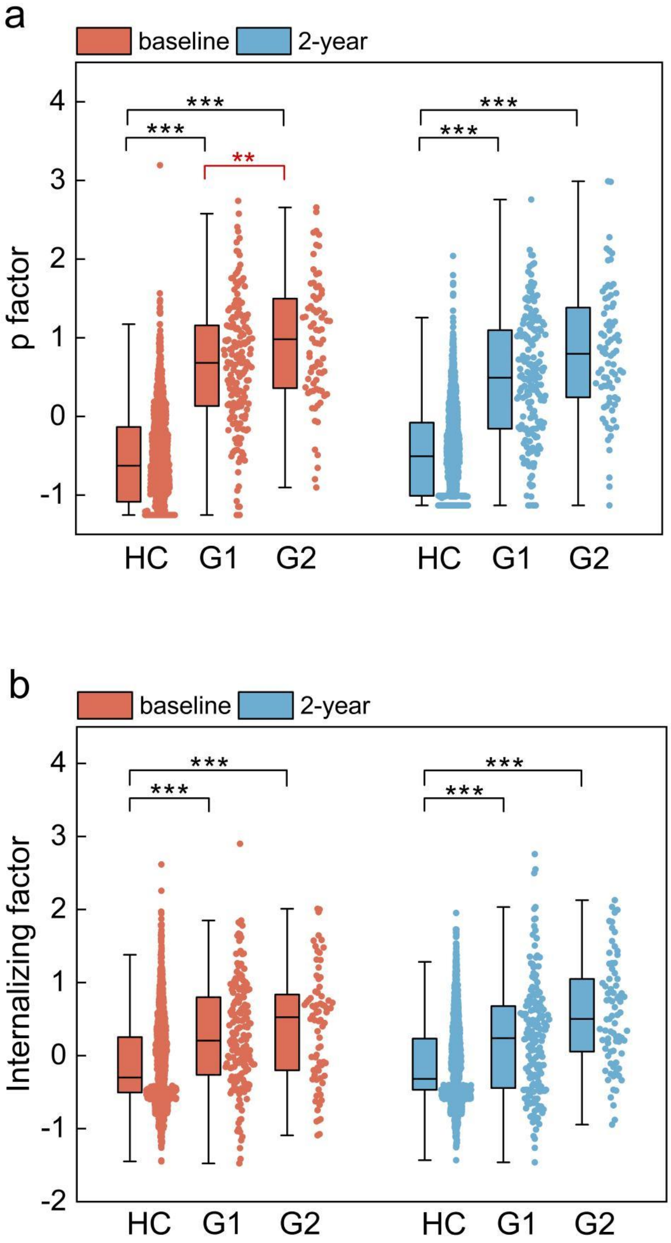

***Note:*** *(a) represents the distribution of the p factor in each group at both baseline and 2-year-follow-up measurements. (b) represents the distribution of the specific internalizing factor in each group at both baseline and 2-year-follow-up measurements. *** = p <.001, ** = p < .01. HC = full Healthy Control group. G1 = Remission group, G2 = Recurrence group.*
